# Differential Gene Expression of Tumors Undergoing Lepidic-Acinar Transition in Lung Adenocarcinoma

**DOI:** 10.1101/2024.03.18.24304449

**Authors:** Ethan N. Okoshi, Shiro Fujita, Kris Lami, Yuka Kitamura, Ryuta Matsuda, Takuro Miyazaki, Keitaro Matsumoto, Takeshi Nagayasu, Junya Fukuoka

## Abstract

Lung adenocarcinoma is the most frequent subtype of thoracic malignancy, which is itself the largest contributor to cancer mortality. The lepidic subtype is a non-invasive tumor morphology, whereas the acinar subtype represents one of the invasive morphologies. This study investigates the transition from a non-invasive to an invasive subtype in the context of lung adenocarcinoma.

Patients with pathologically confirmed mixed subtype tumors consented to analysis of RNA-seq data extracted from each subtype area separately.

The study included 17 patients with tumors found to exhibit a lepidic-acinar transition. 87 genes were found to be differentially expressed between the lepidic and acinar subtypes, with 44 genes significantly upregulated in lepidic samples, and 43 genes significantly upregulated in acinar samples. Gene ontology analysis showed that many of the genes upregulated in the acinar subtype were related to immune response. Immune deconvolution analysis showed that there was a significantly higher proportion of M1 macrophages and total B cells in acinar areas. Immunohistochemistry showed that B cells were mainly localized to tertiary lymphoid structures in the tumor area.

This is the first study to investigate the molecular features of mixed subtype lepidic-acinar transitional tumors. Immunological dynamics are presumed to be involved in this transition from lepidic to acinar subtype. Further research should be conducted to elucidate the progression of disease from non-invasive to invasive morphologies.

## INTRODUCTION

Lung cancer is the largest contributor to cancer mortality and is responsible for over 400,000 deaths per year worldwide.^1,2^ Nearly all lung cancers are carcinomas, the most frequent histologic subtype of which is adenocarcinoma, and its incidence rate is increasing despite public health outreach efforts to limit tobacco usage.^1,3^ Most lung cancers which occur in persons who have never smoked fall within this category– overall, lung adenocarcinoma (LUAD) accounts for 40% of all lung cancers diagnosed in the US.^3^

LUAD is characterized by its origin in the peripheral lung tissue and glandular structure. LUAD is itself subcategorized into five main histologic subtypes: lepidic, acinar, papillary, micropapillary, and solid. Other subtypes are also described, such as invasive mucinous adenocarcinoma, colloid adenocarcinoma, fetal adenocarcinoma, and enteric-type adenocarcinoma.^4^ This distinction was based primarily on histologic characteristics, but research has demonstrated that molecular^5,6^ and clinical^7^ differences between these subtypes can be appreciated as well. In fact, histologic subtype has been shown to significantly correlate with prognostic outcome, with lepidic-predominant tumors (low-grade) having higher 5-year survival rates than acinar (mid-grade), or micropapillary (high-grade) tumors.^8–11^

In clinical practice, there is often a high degree of morphological heterogeneity within a single LUAD, and guidelines generally recommend pathologists to describe the percentage of the specimen area that each subtype comprises.^4,12^ The underlying mechanism of histologic transition from one subtype to another remains to be clarified. Since subtypes of varying grades are regularly seen within a single lesion, it is presumed that differences in protein expression, rather than gene mutations, are more responsible for this histologic transition.^13,14^ However, as the histologic subtyping of LUAD is a relatively recent development, there yet remains a lack of knowledge and clinical experience in this field, and research involving histopathology is fundamentally challenging due to the possibility of conflicting opinions among pathologists.^15,16^ Molecular pathology research is further hindered by the relatively low quality of formalin fixed, paraffin embedded (FFPE)-derived DNA or RNA, as FFPE samples are the standard for histopathological evaluation. Molecular research on the histologic transition between two subtypes necessitates multi-lesion sequencing, which is often labor intensive.

This study seeks to investigate the molecular features of the histologic transition between LUAD subtypes with particular focus on the lepidic-acinar transition. We conducted paired next-generation sequencing of low- and high-grade areas within a single lesion in a cohort of 67 surgically resected patients and compared the molecular signatures of the pairs. We hope that by elucidating the mechanism behind disease progression to a more aggressive pattern, we can spur further research into prognostically significant biomarkers or novel treatment strategies.

## MATERIALS AND METHODS

### PATIENT POPULATION

A cohort of 283 patients with surgically resected primary LUAD from 2009-2015 was retrospectively collected at two institutions (Nagasaki University Hospital, Nagasaki, Japan and Kameda Medical Center, Kamogawa, Japan). All patients had primary solitary LUAD, and patients with mixed histologic findings (e.g., adenocarcinoma with other lung cancer types), metastatic lung tumors, multiple resected lesions, or double lung primary adenocarcinoma were excluded from the study. Surgically resected tissue samples were scanned with a digital slide scanner (20x magnification; Aperio Scanscope CS2, Leica Biosystems, USA), and subtype histology was evaluated via an AI-assisted workflow trained on diagnoses from a panel of expert pathologists,^17^ under supervision by a pulmonary pathologist. The workflow included two deep-learning classification models training on labeling data from 18 expert pathologists. We reviewed 243 patients for which the AI-assisted analysis results were consistent with our visual evaluation and two subtypes were detected. 47 patients with cleanly separated lepidic-acinar subtype transition histology were selected out of this cohort for RNA-seq analysis. Patients in which the subtypes were highly mixed, one subtype was observed at a very low rate, or patients with underlying interstitial lung disease were excluded. This process left 18 patients. RNA was extracted from each subtype area separately and sequenced. In order to confirm the shared lineage of the lepidic and acinar areas present in the surgical sample, we analyzed each sample’s driver gene mutation profile to see if it matched its pair. 17 of the 18 patients had matching driver gene mutation profiles between the sample pairs, and the remaining 1 patient was excluded from the study. We then proceeded with analysis with this cohort of 17 patients with a malignancy that was undergoing lepidic-acinar transition.

All procedures were performed in compliance with relevant laws and institutional guidelines in accordance with the Declaration of Helsinki and have been approved by the Medical Research Ethics Committee of Tokyo Medical and Dental University (M2021-315; July 26^th^, 2022).

### NEXT-GENERATION SEQUENCING

For each patient, 1-3 sections were stained with hematoxylin and eosin (H&E), areas consisting of a given subtype were marked by a pathologist, then unstained FFPE slides were macrodissected manually using a scalpel. RNA was extracted from the macrodissected material using the Qiagen RNeasy FFPE kit (Qiagen, Hilden, Germany), and sequenced using the NEBNext Ultra II Directional RNA Library prep for Illumina kit (New England Biolabs, Ipswich, Massachusetts) and Illumina next-generation sequencers (Illumina, San Diego, California), according to the manufacturers’ instructions.

### BIOINFORMATICS

Raw RNA reads were trimmed using Trimmomatic (v0.39)^18^ and read quality control was performed with FastQC (v0.11.9)^19^ and MultiQC (v1.14).^20^ Trimmed RNA-seq data was then analyzed using CLC Genomics Workbench (v 23.0.4, Qiagen, Hilden, Germany) for read mapping and variant calling, STAR-Fusion^21^ for fusion gene detection, and the Trinity CTAT splicing module for cancer splicing aberrations.

Differential gene expression was analyzed using the RaNA-seq platform.^22^ Results obtained from RaNA-seq were confirmed using the Metascape platform via Gene Ontology (GO) and functional enrichment analysis.^23^ Normal tissue expression data was obtained from the Human Protein Atlas (Human Protein Atlas, proteinatlas.org). Immune cell deconvolution was performed using CIBERSORT-x.^24^

## RESULTS

### COHORT

The method of patient selection for this study is presented as a study flow diagram (**Figure 1A**). Of the 243-patient cohort, the lepidic-acinar transition was the most common mix of subtypes with 47 patients, tied with patients showing acinar-papillary transition (**Figure 1B**). The next most common transition pairing was acinar-micropapillary in 43 patients, followed by acinar-solid in 24 patients. The remaining pairs were seen in 20 or less patients each. For the purpose of this study, we focused on patients displaying a lepidic-acinar transition pattern.

**Figure 1.**
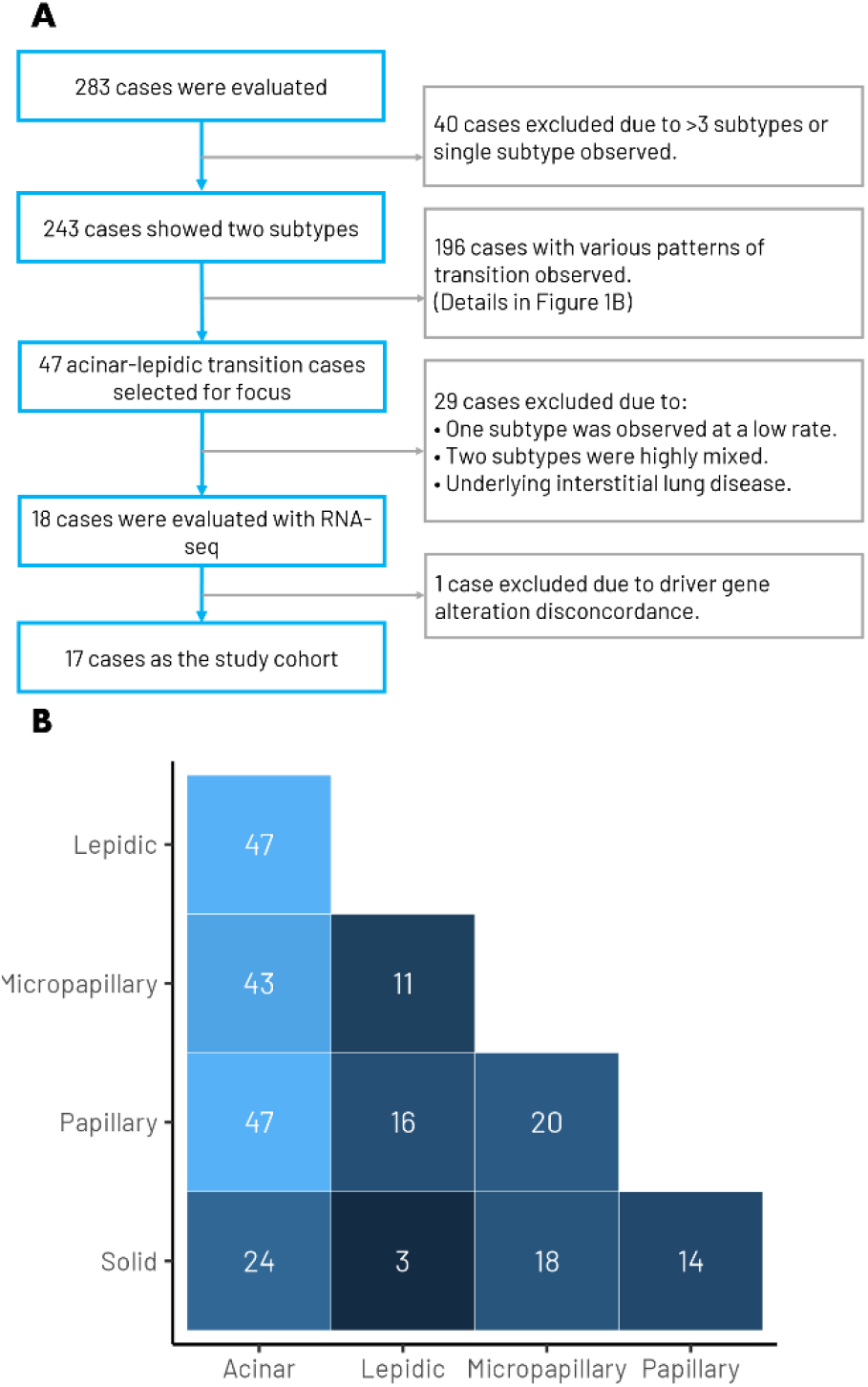
Study design and cohort composition. A) Study cohort design flow diagram. B) Distribution of adenocarcinoma subtype pairs in a 243-patient cohort. Heatmap shows the number of patients for each subtype pairing. Lighter color indicates higher frequency, darker color indicates lower frequency. Patients with malignancies showing lepidic-acinar transition were the most common, along with patients with papillary-acinar transition with 47 patients each. The study cohort was ultimately reduced to 17 patients after clinical and histological evaluation.

17 patients with malignancies exhibiting lepidic-acinar transition were selected based on criteria such as the spatial distribution and relative percentage of each subtype. Clinical data for the 17 selected patients is presented in **Table 1**.

**Table 1.** Patient cohort characteristics. Values for continuous variables are presented as median (IQR), and categorical variables are presented as counts and frequencies as percents. STAS, spread through air spaces.

| Characteristic | N = 17<sup>1</sup> |
| --- | --- |
| Age | 67 (60, 74) |
| Sex |  |
| Female | 10 (59%) |
| Male | 7 (41%) |
| Smoking History |  |
| Ever | 5 (29%) |
| Never or light | 12 (71%) |
| T Stage |  |
| pT1a | 6 (35%) |
| pT1b | 5 (29%) |
| pT1c | 2 (12%) |
| pT1mi | 2 (12%) |
| pT2a | 1 (5.9%) |
| pTis | 1 (5.9%) |
| N Stage |  |
| N0 | 16 (94%) |
| N1a | 1 (5.9%) |
| Stage |  |
| 0 | 1 (5.9%) |
| IA1 | 8 (47%) |
| IA2 | 4 (24%) |
| IA3 | 2 (12%) |
| IB | 1 (5.9%) |
| IIB | 1 (5.9%) |
| STAS | 3 (18%) |
| Lymphatic Invasion |  |
| absent | 15 (88%) |
| present | 2 (12%) |
| Vascular Invasion |  |
| absent | 17 (100%) |

<sup>1</sup>Median (IQR); n (%)

### MUTATIONAL ANALYSIS

In order to confirm the shared lineage of the two tumor subtype areas for each pair, we conducted driver gene mutation analysis on the RNA-seq data. The analysis of the sample pairs showed that all pairs except one showed concordance in their driver gene mutation profiles. The one pair which did not show concordance was excluded from the study **(Supplementary Table 1)**. We observed four pairs expressing KRAS-G12C, three pairs expressing EGFR-L858R, and 10 pairs expressing a wild-type driver gene profile.

### DIFFERENTIAL EXPRESSION

RNA was extracted from macrodissected FFPE specimens, which were annotated by a pathologist to indicate areas of lepidic or acinar pattern. Differential expression analysis was applied to estimate the differential expression between subtype-specific samples of the same tumor. Of 29,264 genes mapped to a reference genome, 87 genes were found to be significantly differently expressed. 44 of those genes had relatively high expression in lepidic components, and 43 had relatively high expression in acinar components (**Figure 2A-C**).

**Figure 2.**
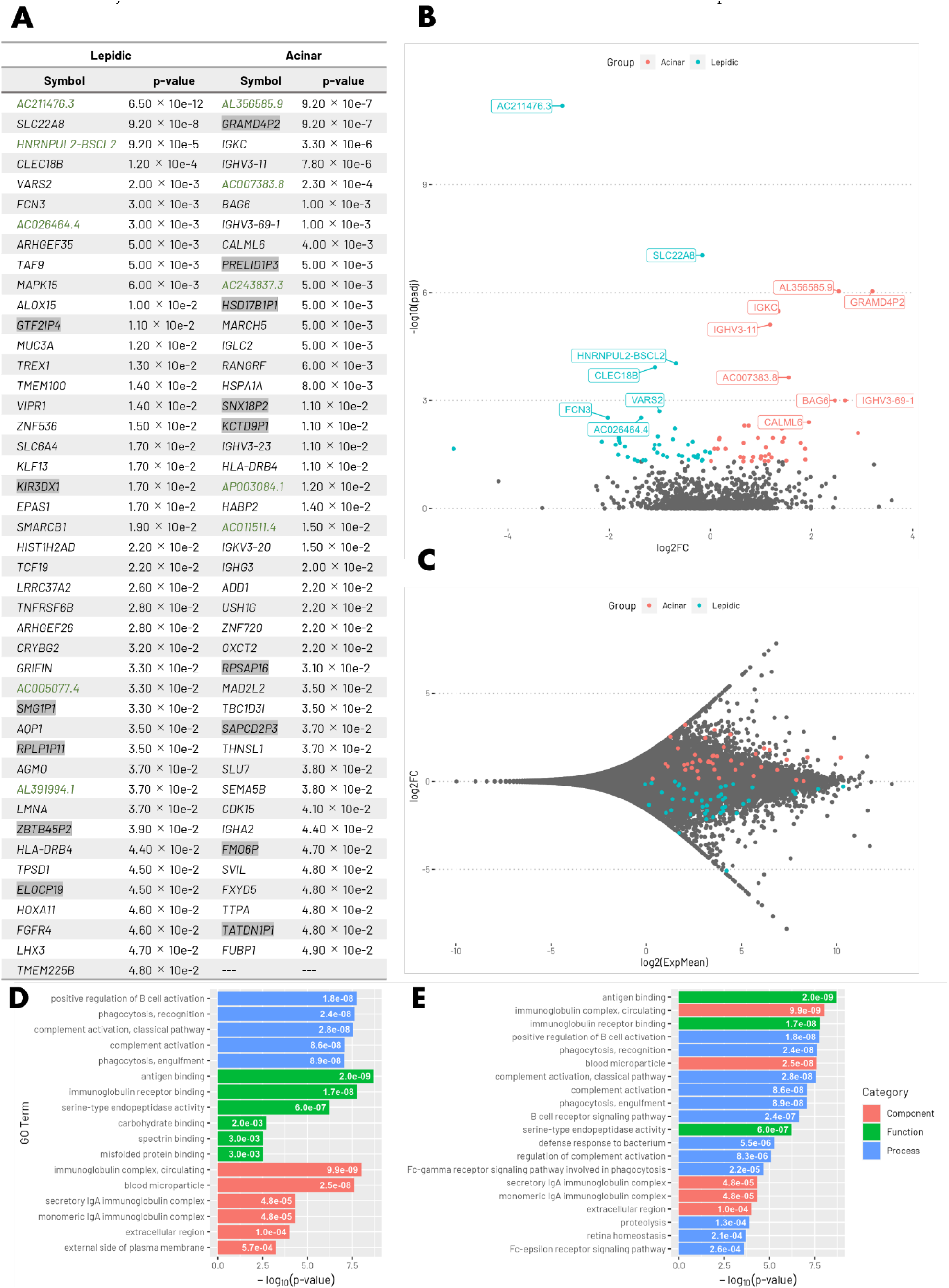
Differential expression and gene set enrichment analysis of RNA-seq data from a Lepidic-Acinar LUAD transition cohort. A) Table showing the significantly differentially expressed genes (DEGs) favoring lepidic and acinar expression profiles respectively. Pseudogenes are highlighted in gray. Non-coding RNA genes are shown in green text. The significance level of the p value was set at 0.05 (results of p = 0.05 were excluded). B) Volcano and C) MA plot visually representing the differential expression of gene transcripts in our dataset. DEGs are colored according to whether they are more highly expressed in lepidic or acinar samples. Genes at p < 0.005 are labeled by name in B. D) bar plot showing the top 5 differentially expressed pathways by category and E) top 20 differentially expressed pathways overall resultant from Gene Ontology (GO) over-representation analysis. Numbers inside the area of the bars are p-values.

Next, we performed over-representation and gene set enrichment (GSEA) analyses. The top 20 resulting categories are presented in **Figure 2D-E**. Most of the gene ontology terms resulting from this analysis are related to immune activity. Antigen binding was found to be the most significantly enriched term, followed by terms related to the immunoglobulin complex and immunoglobulin receptor binding. 13 of the top 20 terms were related to immune function.

In order to confirm the previous results, we repeated the gene ontology analysis on a different bioinformatics platform: Metascape. Once again, the most overexpressed pathways in the acinar subtype correspond to immune-related reaction and cellular activities (**Figure 3**).

**Figure 3.**
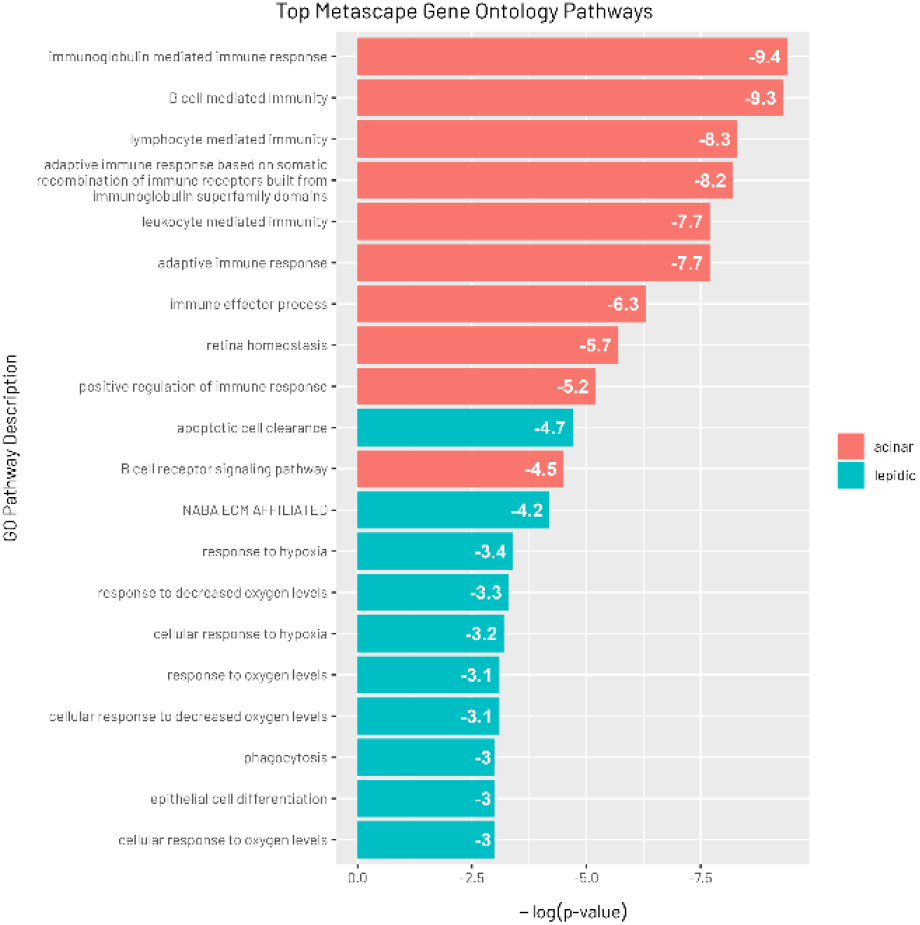
Top 10 represented gene ontology pathways from lepidic and acinar overexpressed gene sets. The Metascape platform was used to analyze the top 44 significantly overrepresented genes from the lepidic group and the top 43 significantly overrepresented genes from the acinar subgroup as described in figure 3A. The Metascape results indicate the most significantly enriched gene ontology pathways based on these lists of genes. The top 10 genes from each group are shown in this figure.

We then examined the context of our genes of interest in normal tissue **(Figure 4**). According to expression signatures from the Human Protein Atlas, genes relatively highly expressed in the lepidic component are most present in healthy lungs, whereas relatively highly expressed genes in the acinar component are seen in lymphoid and intestinal tissue.

**Figure 4.**
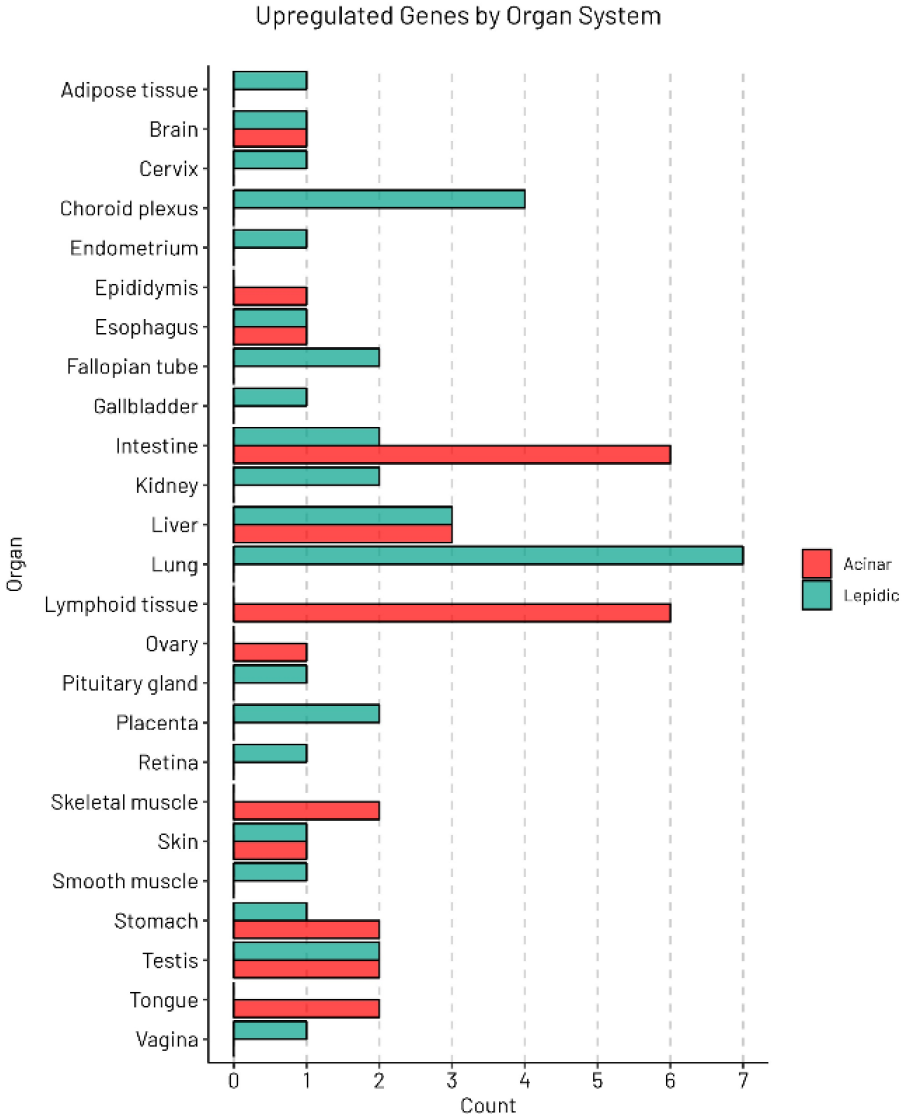
Expression analysis using the Human Protein Atlas. The count of number of upregulated genes is plotted by associated organ system. Genes highly expressed in the lepidic component are related to healthy lung, whereas genes highly expressed in the acinar component are seen most in the lymphoid tissue and the intestine.

### IMMUNE DECONVOLUTION

Next, we estimated the profile of infiltrating immune cells from the bulk tumor expression data using CIBERSORT-x, and compared the relative compositions of 22 immune cells in our samples (**Figure 5**). Of note, a significant difference was observed between lepidic and acinar immune signatures in M1 macrophages, along with borderline significance for CD8+ T cells (**Figure 5C**). M1 macrophages were found to be in higher proportion than other immune cells in the acinar subtype, whereas CD8+ T cells were more relatively common in the lepidic subtype (**Supplemental Figure 2**). When pooling cell types into broader cell type categories, we found that B cells and CD4+ T cells had significantly different fractions between acinar and lepidic groups **(Figure 5C)**.

**Figure 5.**
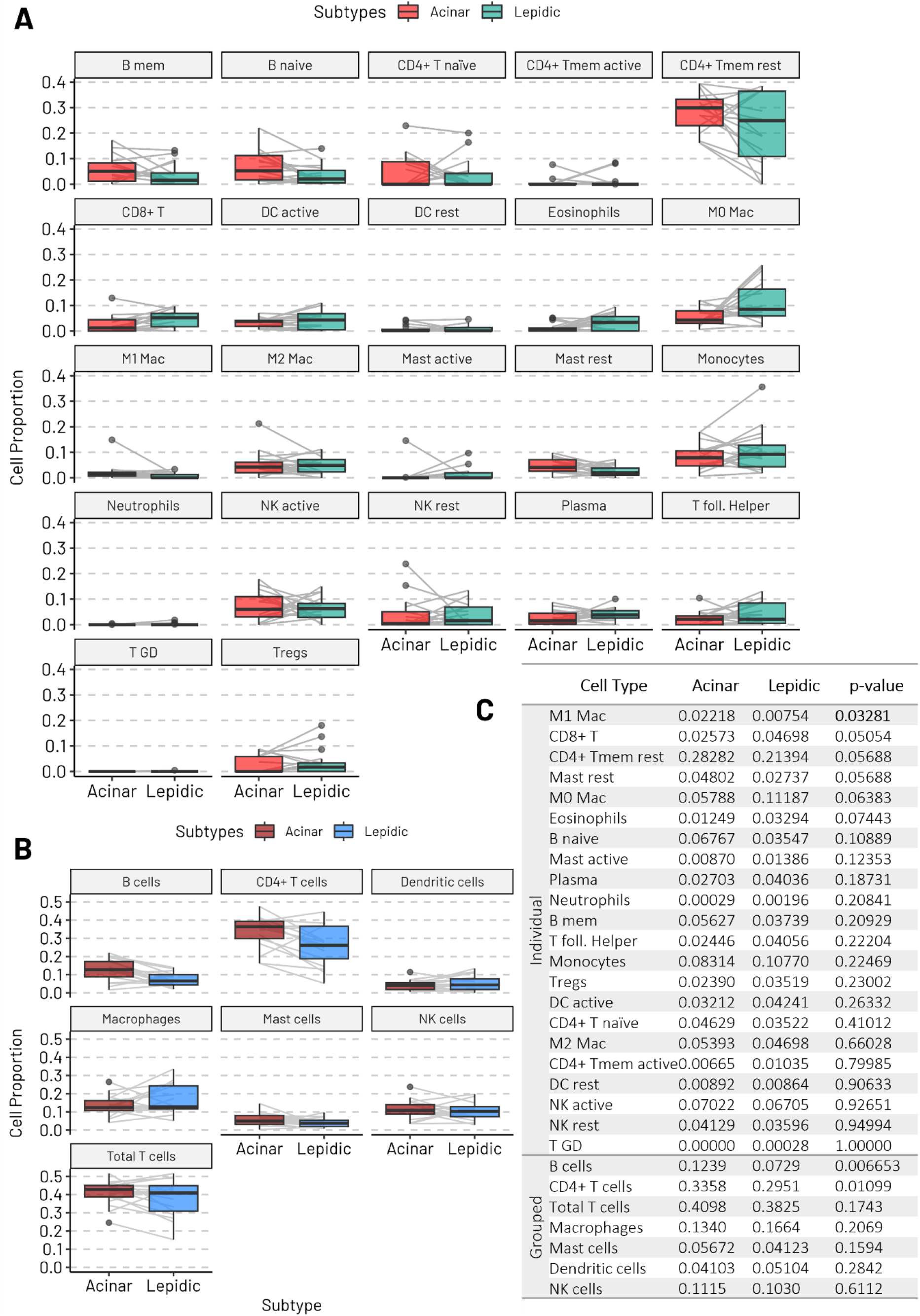
Comparison of immune cell types in acinar vs. lepidic subtype areas. A) Paired box plots showing cell proportions of acinar vs. lepidic samples separated by individual cell types and B) grouped cell types. C) Tabulated data showing values for individual and grouped analysis. Acinar and Lepidic columns show the mean of all samples. P-values were calculated using a two sided pairwise Wilcoxon signed rank test. M1 Mac, M1 macrophages; CD8+ T, CD8+ T cells; CD4+ Tmem rest, resting memory CD4+ T cells; Mast rest, resting Mast cells; M0 Mac, M0 macrophages; B naïve, naïve B cells; Mast active, active Mast cells; Plasma, plasma cells; B mem, memory B cells; T foll. Helper, T follicular helper cells, Tregs, regulatory T cells; DC active, active dendritic cells; CD4+ T naïve, naïve CD4+ T cells; M2 Mac, M2 macrophages; CD4+ Tmem active, active CD4+ memory T cells; DC rest, resting dendritic cells; NK active, active natural killer cells; NK rest, resting natural killer cells; T GD, gamma delta T cells.

We then performed an immunohistochemistry (IHC) assay to identify the location of B cells in the tissue. IHC showed that B cells were mainly localized to tertiary lymphoid structures (TLS) and were present in both lepidic and acinar areas (**Figure 6**).

**Figure 6.**
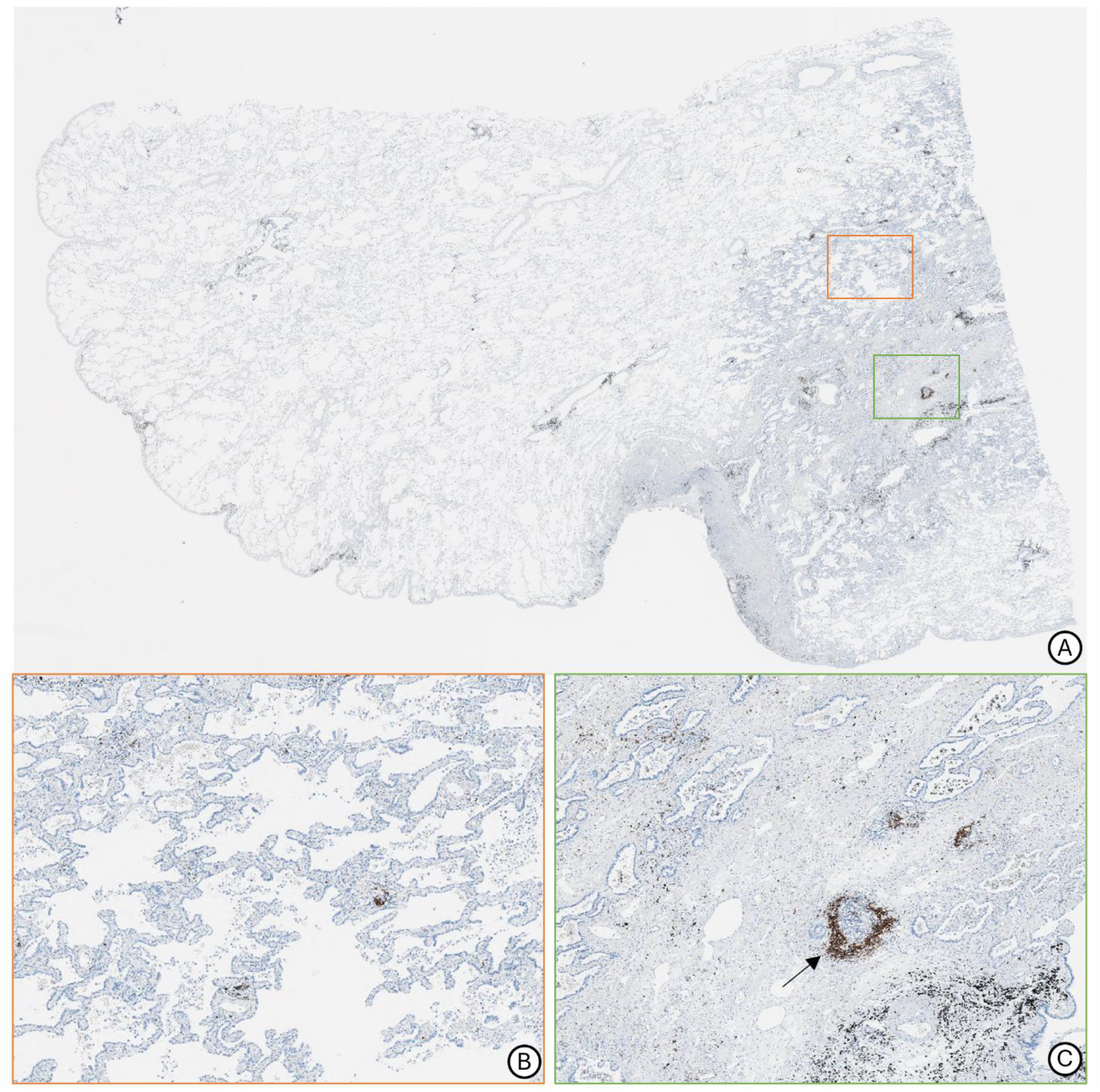
Representative images of CD20 immunohistochemistry staining. A) Overall view of tissue sample with orange and green boxes representing detail areas B and C respectively. B) Detail view of lepidic subtype area with small amounts of B cells scattered nonspecifically. C) Detail view of acinar subtype area. Arrow pointing towards tertiary lymphoid structure with localization of B cells.

## DISCUSSION

This study compared the expression signatures of lepidic and acinar subtypes sampled from within the same LUAD lesion. The gene expression of the acinar group saw significant upregulation in immune system-related genes, specifically those relating to immunoglobulins. Conversely, the top three differentially expressed ontology clusters in the lepidic group were apoptotic cell clearance, extracellular matrix proteins, and response to hypoxia. Computational deconvolution revealed that M1 macrophages, B cells, and CD4+ T cells were significantly more abundant in the acinar group than in the lepidic. Immunostaining confirmed that B cells were localized to TLS within the tumor tissue.

We found that M1 macrophages with intrinsic anti-tumor properties were more common in acinar areas, which is a higher-grade histologic subtype. The tumor microenvironment (TME) includes several types of cells and molecules, including immune cells, blood vessel cells, and connective tissue cells.^25^ In the TME, macrophages, T cells, natural killer cells, and dendritic cells are said to play a major role, and among these immune cells, macrophages account for a large proportion. Tumor assisted macrophages (TAMs) differentiate from macrophages through various factors present in the TME. It is also known that macrophage polarization produces two functionally different phenotypes, M1 and M2.^26^ M1 macrophages produce ROS and induce the production of many inflammatory factors such as TNF-a, IL-6 and IL-1B, and play important roles in pathophysiologic processes such as resistance to tumor cells, killing pathogenic microorganisms, and anti-inflammatory responses. M2 macrophages exert immune suppression and promote tumor progression by secreting fibroblast growth factor (FGF), matrix metalloproteinase, and interleukins.^27,28^ It should be noted that changes in M1 and M2 are reversible processes, and each phenotype is identifiable via surface antigen markers.

One possible reason for the significantly greater number of M1 macrophages in the acinar subtype is the hypoxic environment. Macrophage polarization is a complex process co-regulated by multiple signaling molecules and their signaling pathways including JAK/STAT, PI3K/AKT, JNK, and Notch.^29^ Metabolic change also plays an important role in polarization. M1 macrophages mainly utilize glycolysis to meet biosynthesis and energy needs.^26^ The glycolysis pathway is an anaerobic reaction that does not require oxygen. As the lepidic to acinar transition is inevitably accompanied with a loss of air space, this process may lead to hypoxic conditions in the TME, which induces macrophage differentiation to the M1 state. In addition, the expression level of OXCT2 was found to be significantly higher in the acinar subtype than in the lepidic. The protein encoded by this gene is an important enzyme in ketone body catabolism. Tissues that normally obtain energy using glucose use ketone bodies such as β-hydroxybutyrate and acetoacetate when glucose is unavailable (such as during starvation). The use of ketone bodies suggests that tumor cells in acinar tissues are more dependent on glycolysis than those in lepidic tissues, and the increased expression of OXCT2 reflects the hypoxic state of the acinar tissue.

There are several reports that non-small cell lung cancer (NSCLC) tissue contains more B cells than normal tissue, however, there is no consistent view as to whether there is a difference between B cell levels present in acinar and lepidic tissues. Bruno and colleagues demonstrated that tumor-infiltrating B cells were increased in frequency and abundance compared with tumor-adjacent normal tissue in all subtypes of NSCLC, most prominently in adenocarcinomas.^30^ Across the cohort of patients, only B cells showed a significantly higher representation in NSCLC tumors compared to the distal lung.^31^ Different results have been reported regarding the difference in B cell numbers between lepidic and acinar subtypes. There is one study which examined the differences in the composition of infiltrating immune cells by subtype using immunostaining. They reported that acinar tissues had more B cells than lepidic.^32^ However, this study used acinar and lepidic tissues sourced from different lesions. Another group estimated infiltrating immune cells by computational deconvolution of bulk RNA-seq data, similar to our study. They also showed that acinar tissue was infiltrated by more B cells, although this too was not a comparison within a single lesion.^14^ On the other hand, another group using computational deconvolution reported more B cells in the lepidic subtype, although no statistical significance was found.^11^

We speculate that the reason for the different results among subtypes may be due to differences in the extent to which tertiary lymphoid structures (TLS) were included in the evaluated tissue samples. TLS are de novo structured lymphoid devices that form in non-lymphoid tissue and are known to occur in various solid tumor tissues.^33^ It has been reported that the distribution of TLS within a tumor is associated with the biological grade of the tumor,^34,35^ and some groups have classified TLS themselves into multiple subtypes and reported their relationship to prognosis.^36^ There are several methods to evaluate TLS, including immunostaining and deconvolution of flow cytometry and bulk RNA-seq data, but technical issues, such as differences in the number of TLS depending on the angle of sectioning, limit the ability to establish a gold standard for evaluation.^37^ In our study, we compared different subtypes within a single lesion, and we believe that the results obtained here are more rigorous than those obtained between different lesions. Using this methodology, we established that B cells are localized in the TLS of acinar and lepidic subtypes of lung adenocarcinoma, and that more B cells are present in acinar tissues, which confirms prior reports.^14^

One of our results, the high infiltration rate of B cells in acinar tissues, cannot be interpreted definitively as a response to hypoxia. Regarding the association between an increase in the number of B cells infiltrating into tumor tissue and a hypoxic environment, several reports indicate that a hypoxic environment activates regulatory B cells. Analysis of pancreatic cancer in mice and humans has shown that a hypoxic environment induces upregulation of HIF1a, activates regulatory B cells, and increases the number of cells at lesion sites.^38^ However, the present study did not fully examine the type of infiltrating B cells, so the association between the hypoxic environment and the abundance of B cells in the acinar subtype is unclear.

In this study, CD4+ T cells were also significantly more common in the acinar subtype, and in particular, CD8+ T cells and memory resting CD4+ T cells were more common in the acinar subtype, albeit with borderline significance. CD8+ cells are often discussed because the percentage of CD8-positive cell infiltration is associated with response to immune checkpoint inhibitors. Perhaps for this reason, the behavior of CD4+ cells in NSCLC has not been well studied, even though CD4+ lymphocytes are the most common immune cells found in lung adenocarcinoma tissue. Prior reports have shown that resting memory CD4+ T cells are decreased in poorly differentiated subtypes, i.e. acinar.^39^ Another report states that among the five major subtypes, the acinar subtype has the highest number of CD4+ T cells.^14^

We found in our study that the gene *HLA-DRB4* was included in the differentially expressed gene lists of both acinar and lepidic subtypes. This is due to the fact that HLA-DRB4 is highly polymorphic and in our RNA-seq methodology, mapping is performed for each polymorphic site. Thus, this gene is particularly difficult to analyze precisely by short-read-based RNA-seq precisely. Long-read sequencing is recommended for the analysis of this gene family.

The strength of our study is that we selected and analyzed areas of differing LUAD subtypes from a single lesion. To our knowledge, this is the first study to analyze the lepidic-acinar subtype transition using tissue derived from the same lesion. We have molecularly verified that our samples represent a transition from a non-invasive to an invasive subtype by excluding patients in which the driver gene mutation profile of each subtype area did not match. Furthermore, the use of AI for histological type determination eliminates the situation in which judgements differ between evaluators. On the other hand, the limitation of the present study is that only expression analysis was performed, lacking DNA-based mutation analysis. The present study was designed based on previous reports which suggested that changes in tissue subtype were not due to changes in gene mutation patterns, but rather to changes in protein expression status.^13,14^ In addition, since RNA extraction was required after tissue type determination using whole slide image-based AI, formalin fixed paraffin embedded (FFPE) samples were used as the source of genetic material. In the future, analysis by single-cell RNA-seq will provide more extensive and comprehensive information related to changes in gene expression related to subtype transition.

## CONCLUSION

We performed RNA-seq on a single lesion showing subtype transition from lepidic to acinar and identified a group of genes associated with subtype change. We found that there were 44 genes that were significantly expressed in the lepidic component of our samples, and 43 genes that were significantly expressed in the acinar component of our samples. The genes corresponding to the lepidic component are mainly expressed in healthy lungs, and the genes related to the acinar component are normally expressed in lymphoid tissue and intestine. Pathway analysis showed that many of these genes are related to immune response, particularly for genes upregulated in the acinar subtype. Immune deconvolution analysis supported the idea that hypoxia may play a role in this transition.

With advances in sequencing technology and bioinformatic resources, further research is warranted in order to explore the molecular mechanisms behind the transition to more invasive disease in LUAD. The discovery of biomarkers and mutational patterns can lead to improved prognostication and treatment strategies. Extending the analysis of transitions within a single lesion to other subtypes will provide new insights into the progression of this refractory malignant disease and refine strategies against it.

## Author Contributions

Ethan N. Okoshi: Formal analysis, Investigation, Data curation, Writing-Original draft, Visualization. Shiro Fujita: Conceptualization, Methodology, Formal analysis, Writing – Original draft, Supervision. Kris Lami: Methodology, Formal analysis, Investigation, Data curation, Writing – Review and editing. Yuka Kitamura: Investigation, Resources. Ryuta Matsuda: Investigation, Resources. Takuo Miyazaki: Resources. Keitaro Matsumoto: Resources. Takeshi Nagayasu: Resources. Junya Fukuoka: Conceptualization, Methodology, Resources, Writing – Review and editing, Supervision, Project administration, Funding acquisition.

## Ethics Declaration

All procedures were performed in compliance with relevant laws and institutional guidelines in accordance with the Declaration of Helsinki and have been approved by the Medical Research Ethics Committee of Tokyo Medical and Dental University (M2021-315; July 26^th^, 2022). Informed patient consent was obtained.

## Data Availability Statement

Data is available upon reasonable request from the corresponding author.

## SUPPLEMENTAL MATERIALS

**Supplementary Table 1.**
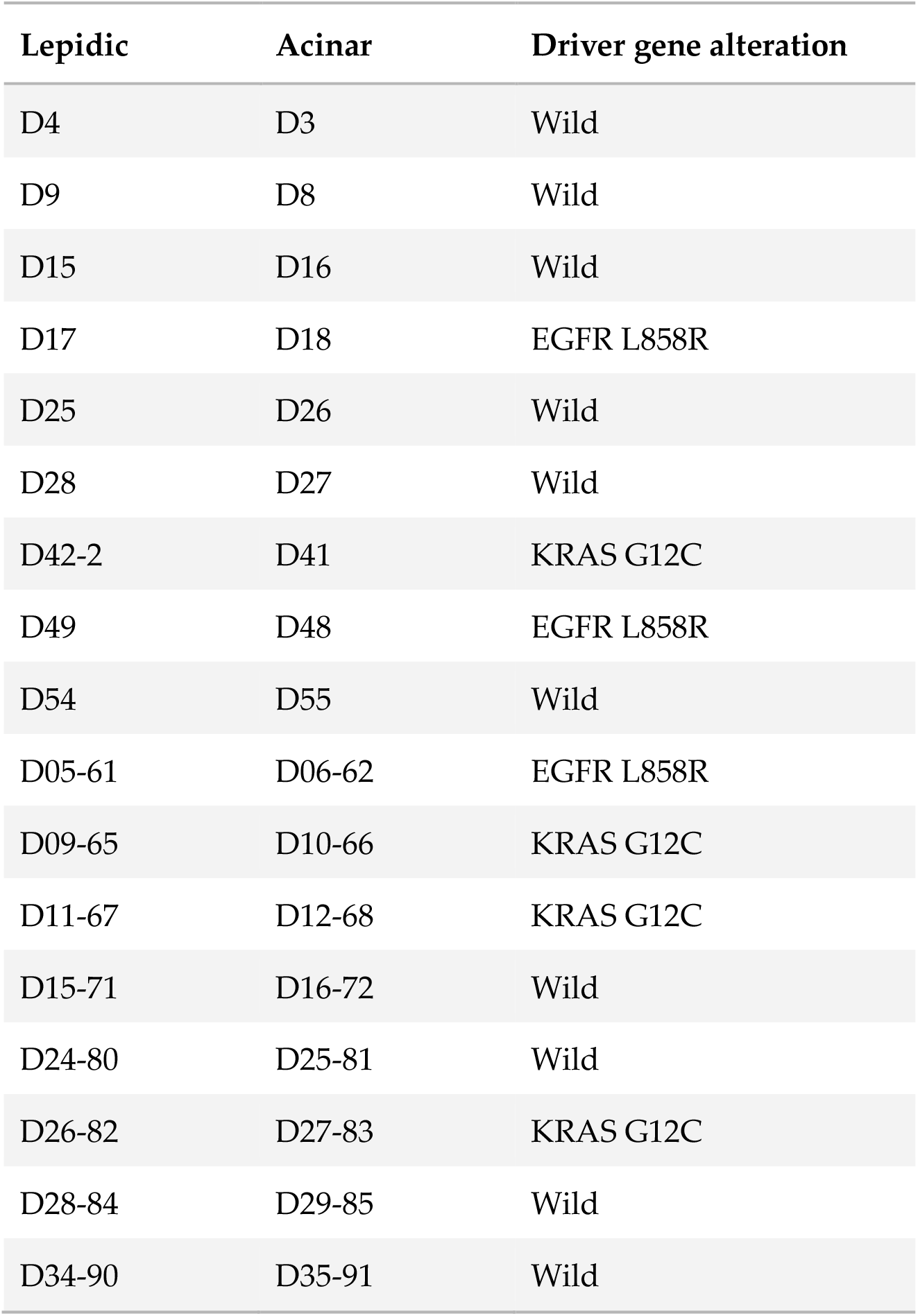
Driver gene mutation analysis of sample pairs. Values in left columns are case identifiers.

| **Lepidic** | **Acinar** | **Driver gene alteration** |
| --- | --- | --- |
| D4 | D3 | Wild |
| D9 | D8 | Wild |
| D15 | D16 | Wild |
| D17 | D18 | EGFR L858R |
| D25 | D26 | Wild |
| D28 | D27 | Wild |
| D42-2 | D41 | KRAS G12C |
| D49 | D48 | EGFR L858R |
| D54 | D55 | Wild |
| D05-61 | D06-62 | EGFR L858R |
| D09-65 | D10-66 | KRAS G12C |
| D11-67 | D12-68 | KRAS G12C |
| D15-71 | D16-72 | Wild |
| D24-80 | D25-81 | Wild |
| D26-82 | D27-83 | KRAS G12C |
| D28-84 | D29-85 | Wild |
| D34-90 | D35-91 | Wild |

**Supplementary Table 2.**
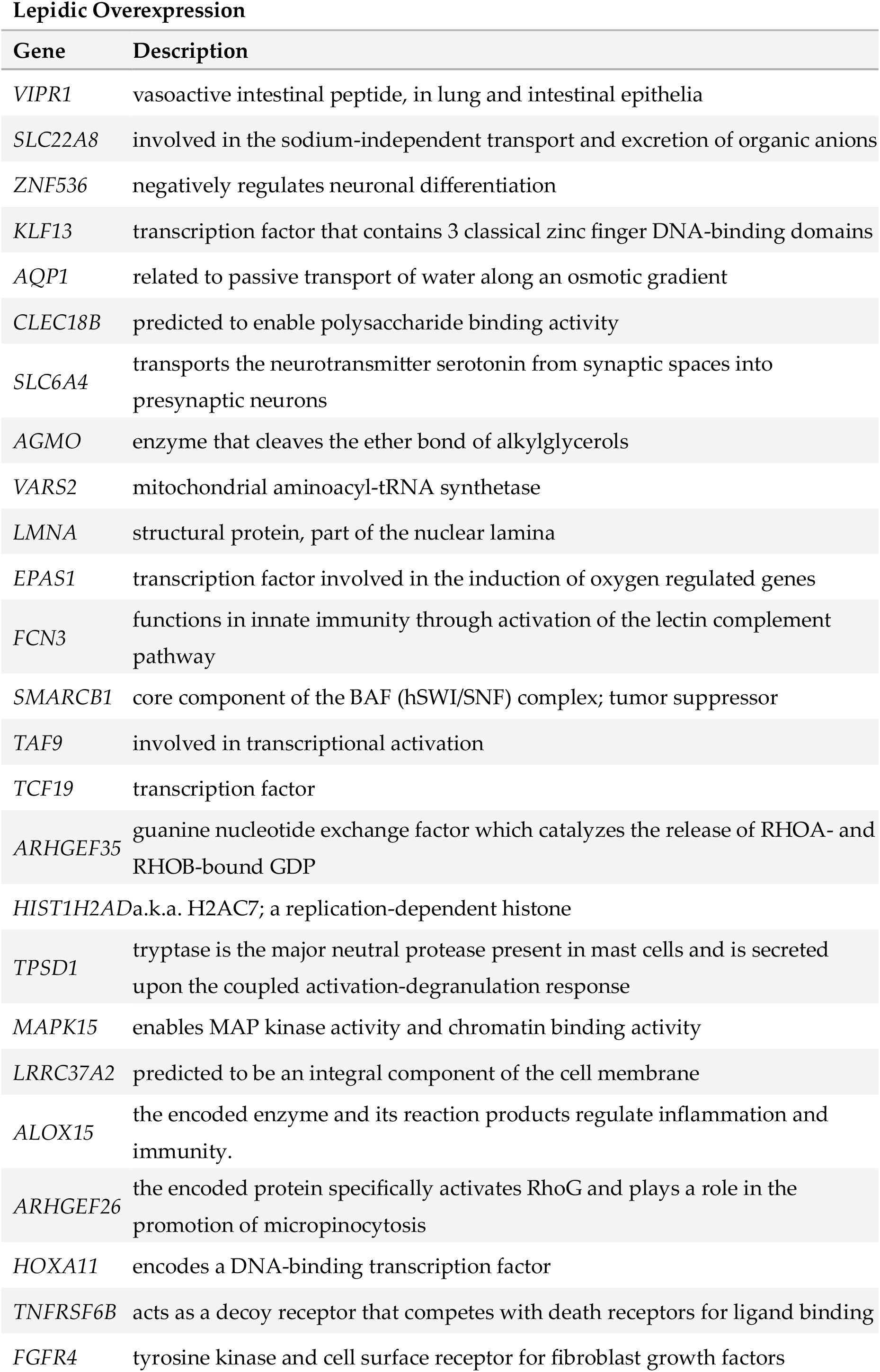
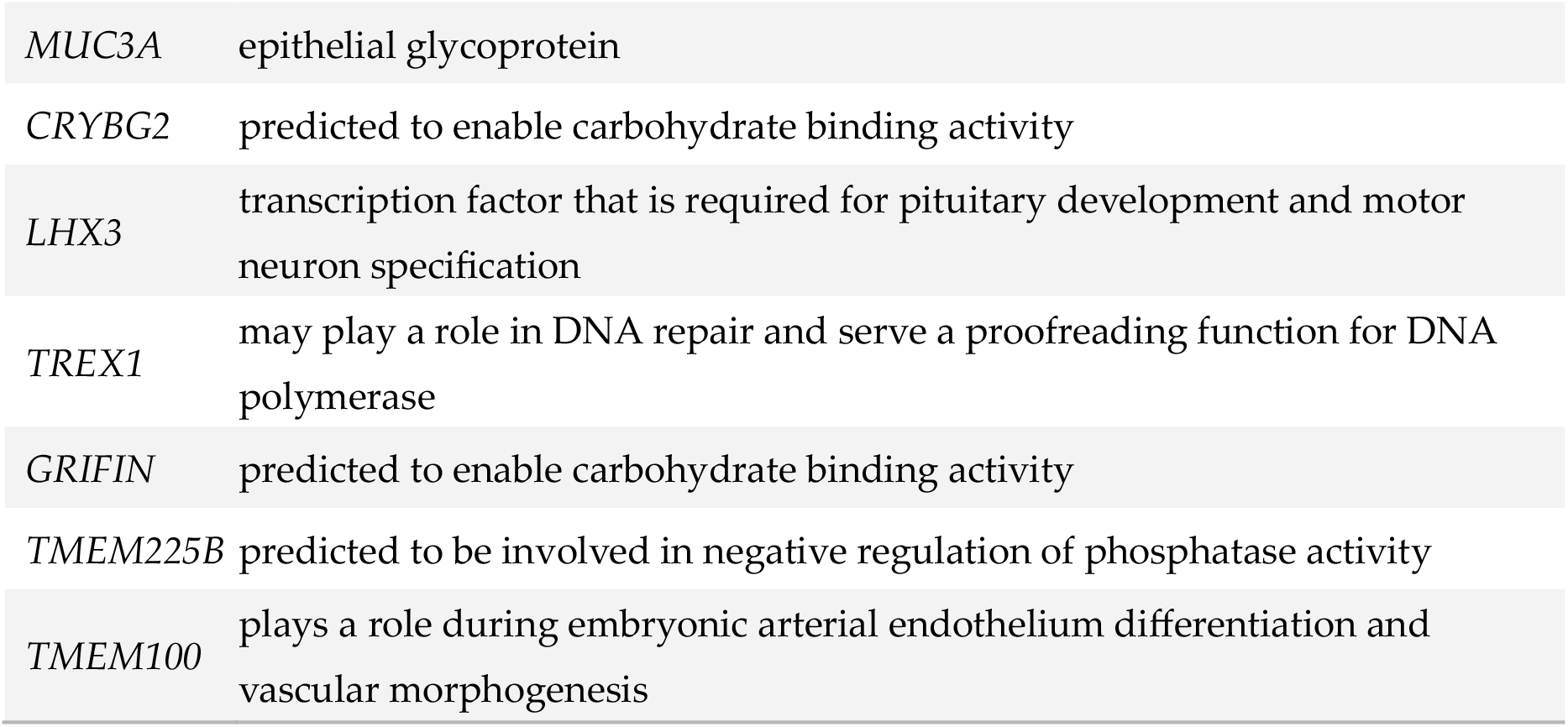
Descriptions of genes upregulated in lepidic areas.

**Supplementary Table 3.**
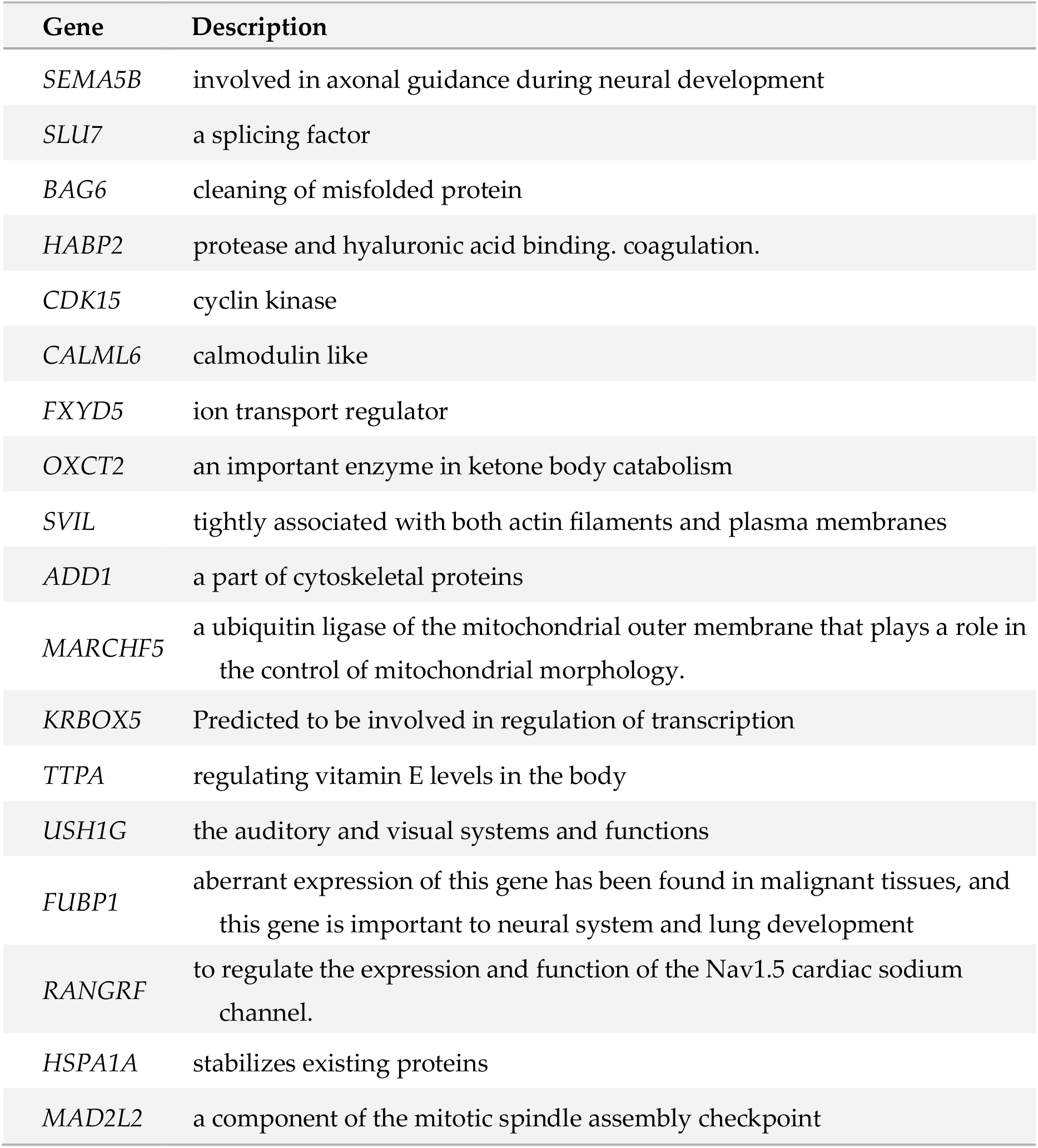
Descriptions of genes upregulated in acinar areas.

**Supplemental Figure 2.**
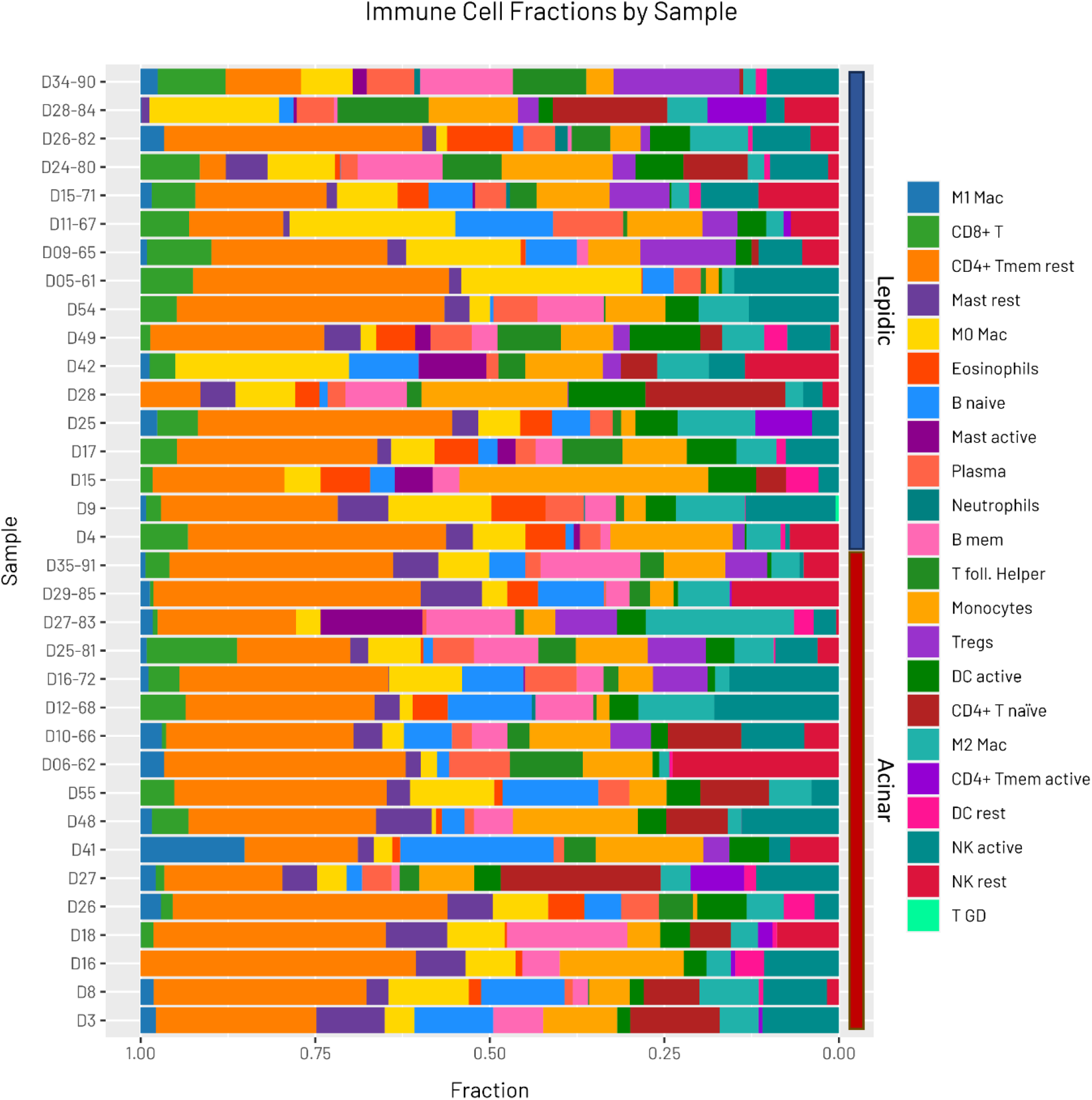
Stacked bar chart showing the relative cell proportions across samples. M1 Mac, M1 macrophages; CD8+ T, CD8+ T cells; CD4+ Tmem rest, resting memory CD4+ T cells; Mast rest, resting Mast cells; M0 Mac, M0 macrophages; B naïve, naïve B cells; Mast active, active Mast cells; Plasma, plasma cells; B mem, memory B cells; T foll. Helper, T follicular helper cells, Tregs, regulatory T cells; DC active, active dendritic cells; CD4+ T naïve, naïve CD4+ T cells; M2 Mac, M2 macrophages; CD4+ Tmem active, active CD4+ memory T cells; DC rest, resting dendritic cells; NK active, active natural killer cells; NK rest, resting natural killer cells; T GD, gamma delta T cells.

